# Waning Effectiveness of the Third Dose of the BNT162b2 mRNA COVID-19 Vaccine

**DOI:** 10.1101/2022.02.25.22271494

**Authors:** Tal Patalon, Yaki Saciuk, Asaf Peretz, Galit Perez, Yoav Lurie, Yasmin Maor, Sivan Gazit

**Affiliations:** Kahn Sagol Maccabi (KSM) Research & Innovation Center, Maccabi Healthcare Services, Tel Aviv, 68125, Israel; Maccabitech Institute for Research and Innovation, Maccabi Healthcare Services, Israel; Internal Medicine COVID-19 Ward, Samson Assuta Ashdod University Hospital, Ashdod Israel; Liver unit, Shaare Zedek City Center Campus, Jerusalem, Israel; Faculty of Medicine, Tel Aviv University, Tel Aviv, Israel; Infectious Disease Unit, Edith Wolfson Medical Centre, Holon, Israel

**Keywords:** COVID-19, SARS-CoV-2, vaccination, waning immunity, booster, BNT162b2

## Abstract

The duration of protection of the third (booster) dose of the BioNTech/Pfizer BNT162b2 mRNA Coronavirus Disease 2019 vaccine has yet to be sufficiently researched, while global discussions around the necessity and effectiveness of a fourth dose are already underway. By conducting a retrospective study implementing a test-negative case-control design, analyzing 546,924 PCR tests performed throughout January 2022 by 389,265 persons who received three doses of the vaccine, we found that the effectiveness in each month since vaccination decreased significantly. Compared to those vaccinated early, on August 2021, relative protection against infection waned from 53.4% a month after vaccination to 16.5% three months after vaccination. These results suggest that there is a significant waning of vaccine effectiveness against the Omicron variant of the third dose of the BNT162b2 vaccine within a few months after administration and should prompt policy discussion as to additional booster doses and vaccine development.

## Introduction

Both the short-term effectiveness of two doses of the BioNTech/Pfizer BNT162b2 mRNA coronavirus disease 2019 (COVID-19) vaccine^1–4^ and the waning of the vaccine-induced immunity have been demonstrated in previous research,^5–9^ though the latter has been mild against severe disease.^10^ These studies, alongside the surge of breakthrough infections with the Delta Variant (B.1.617.2) of the SARS-CoV-2 led policy makers around the globe to administer the third (booster) dose of the vaccine.

In Israel, an early adopter of the two-dose BNT162b2 regimen, a similarly rapid campaign was carried out for the third dose. The booster was first approved on July 30, 2021 for all individuals aged 60 or older who received the second dose at least five months prior, and by August 29, 2021, the campaign was extended to include all individuals aged 12 or older. This swift rollout of the booster facilitated early real-world investigations of its short term effectiveness, largely demonstrating a restored short-term effectiveness^7,9,11,12^.

However, the question of the duration of protection of the third dose and its effectiveness against different variants has yet to be sufficiently researched. Nonetheless, in Israel, the recent surge of infections with the highly transmissible Omicron (B.1.1.529)^13,14^ variant has led to the January 2, 2022 official recommendation to administer a fourth dose of the vaccine to medical personnel and people ages 60 and older,^15^ while expanding the eligibility criteria is currently under discussions.^16^

To this end, we conducted a retrospective analysis examining the association between booster breakthrough infections with the Omicron variant and time-since-vaccination, leveraging data from Maccabi Healthcare Services (MHS), an Israeli health fund that covers 2.5 million lives.

## Methods

### Study population and data sources

The study population included all MHS members aged 16 or older who received at least two doses of the BNT162b2 vaccine by August 1, 2021. Individuals were excluded from the study if they had a positive SARS-CoV-2 polymerase chain reaction (PCR) assay test result, disengaged from MHS for any reason prior to the start of the study period or joined MHS prior to March 2020, hence might have an incomplete COVID-19-related history. Anonymized Electronic Medical Records (EMRs) were retrieved from MHS’ centralized computerized database, a state-mandated, non-for-profit, health fund in Israel which covers 26.7% of the population. This centralized database has been maintained for over three decades, allowing for a comprehensive longitudinal medical follow-up, including demographic data, clinical measurements, outpatient and hospital diagnoses and procedures, medications dispensed, imaging performed and comprehensive laboratory data from a single central laboratory.

### Data extraction

Individual-level demographic data of the study population included sex, age and a coded residential socioeconomic geographical statistical area (GSA), assigned by Israel’s Central Bureau of Statistics, which is the smallest geostatistical unit of the Israeli census (corresponds to neighborhoods) and embodies the socioeconomic status. COVID-19 related data consisted of dates of vaccination and results of any polymerase chain reaction (PCR) tests for SARS-CoV-2, given that all such tests are recorded centrally. COVID-19-related hospitalizations and mortality records were retrieved as well. Captured data also consisted of information on chronic diseases from MHS’ automated registries, including cardiovascular diseases^20^, hypertension, diabetes^21^, chronic kidney disease (CKD)^22^, chronic obstructive pulmonary disease (COPD), obesity (defined as a body mass index of 30 or higher) and immunocompromised conditions.

### Measured Outcomes

We evaluated two SARS-CoV-2-related outcomes. The first was breakthrough infections, defined as a positive documented RT-PCR test 7 days or more after a vaccine dose (i.e., a booster breakthrough infection was defined as a positive test taken 7 or more days after inoculation and not after the fourth dose, if such was administered). The 7-day cutoff was chosen based on previous research the BNT162b2 vaccine effectivenesss.^1,11^ The second outcome referred to a severe disease, defined as a COVID-19-related hospitalization or death occurring within the same time frame. Outcomes were evaluated during the follow-up period of January 1 to January 21, 2022, an Omicron-dominant period in Israel.^13^

The analysis was stratified to different groups, each stratum represented a different time-since-vaccination. The rationale is that if no waning of the booster exists, the rates of breakthrough infection will not depend on the time-from-vaccination. However, if waning of the booster does occur, protection conferred by the third dose will gradually decrease with time-since-vaccination.

### Design and Statistical Analysis

#### Main analysis – test-negative case-control design

Our main analysis consisted of a test-negative case-control design. The test negative design, which has long been used in vaccine effectiveness research, is becoming increasingly common in SARS-CoV-2 vaccine studies, due to its strength in controlling against bias stemming from healthcare seeking behavior.^11,23,24^ In this analysis, we defined cases as PCR-positive persons or persons with a severe manifestation of COVID-19 during the follow-up period (cases were defined for each of the two outcomes separately). Eligible controls were PCR negative individuals who had neither tested positive prior to the date of the positive PCR or the COVID-related hospitalization of their matched caseand whose test was not done after administration of the fourth vaccine, if such was administered. A 1:1 matching was performed based on sex, age group (16 to 29 years, 30 to 39 years, 40 to 49 years, 50 to 59 years, and ≥60 years), GSA, calendrical week of testing, and the month of receipt of the second dose. The first positive PCR test and the first negative PCR test were the only tests included for each case and control, respectively. All negative PCRs for cases were excluded from the study, therefore an MHS member was either a case or a control, not both.^6^

The analysis sought to estimate the reduction in the odds of a positive outcome at different time points following the receipt of the booster dose. Therefore, we stratified the analysis for each month-since-vaccination, where in each stratum matched pairs included only cases that met that specific month since vaccination or the reference group of August 2021vaccinees (hence, the number of matched pairs differed between time points).^6^ The vaccination status (and time from inoculation) was determined at the time of the PCR test (for the first outcome assessing breakthrough infections) or the COVID-19 hospitalization (for the second outcome assessing severe disease).

A conditional logistic regression model was fit to the data, accounting for the matching. The vaccine effectiveness (VE) of the booster in each month, compared to the first month of eligibility for the booster^17^ (August 2021) was calculated as 100%*[1-(Odds Ratio)] for each of the month-since-vaccination categories. To address potential confounders, we adjusted for underlying comorbidities, including obesity, cardiovascular diseases, diabetes, hypertension, chronic kidney disease, COPD and immunosuppression conditions.

#### Additional Analyses

We carried out three additional analyses. First, as a sensitivity analysis for the occurrence of a booster breakthrough infection, we repeated the test-negative analysis, this time unmatched allowing individuals to contribute multiple negative tests, but excluding them once they tested positive (further details can be found in Patalon et al).^11^ In order to account for repeated sample collection from the same individual, a Generalized Estimating Equation (GEE) logistic regression model was fit to the data.^25 25^ Adjustment included demographics and the same comorbidities as the main analysis, in addition to a series of dummy variables representing the calendar week in which the PCR test was performed to address exposure. We assumed an exchangeable correlation structure. In our second sensitivity analysis, we reran the main analysis while including tests that were performed 0 to 6 days after the administration of the fourth dose of the vaccine. The rationale was that if we consider ‘fully vaccinated’ individuals as those 7 days or more after receiving a fourth dose (similarly to the booster), tests performed within that period should not be excluded. This also allowed us to mitigate a potential selection bias of excluding tests of early vaccinees.

Finally, we performed the main analysis again while changing the reference group; instead of the earliest booster vaccinees (i.e., those vaccinated in August 2021), we compared each stratum of time-since-booster to those unvaccinated with the booster, namely persons who received *only* two doses of the vaccine. The advantage of such an analysis is the ability to assess whether the *marginal* effectiveness of the booster over the second dose decreases with time. However, as by the outcome period of January 2022 most of Israel’s population had already received the booster, the reference group of those unvaccinated is admittedly prone to bias, as has been discussed in previous papers.^11^ Therefore, in light of this concern, we chose to present this analysis as a supplementary one.

All analyses were performed using R Studio version 3.6 with the MatchIt, gee and survival packages.

## Data Availability

According to the Israel Ministry of Health regulations, individual-level data cannot be shared openly. Specific requests for remote access to de-identified community-level data should be referred to KSM, Maccabi Healthcare Services Research and Innovation Center.

## Ethics declaration

This study was approved by the MHS (Maccabi Healthcare Services) Institutional Review Board. Due to the retrospective design of the study, informed consent was waived by the IRB, as all identifying details of the participants were removed before computational analysis.

## Code availability statement

Specific requests for remote access to the code used for data analysis should be referred to KSM, Maccabi Healthcare Services Research and Innovation Center.

## Results

546,924 PCR tests by 389,265 MHS members were performed during the outcome period of January 1 to January 21, 2022. 53,486 tests (performed by 38,145 individuals) were excluded from the main analysis, as they were conducted after the administration of the fourth dose (see additional analyses). Population characteristics can be found in **Table 1**. Overall, those vaccinated early were more chronically ill, likely correlating with earlier compliance to vaccination by the older population, emphasizing the need for adjustment.

**Table 1.** Population characteristics by exposure groups in tested population

|  | Exposure Groups/Booster period |  |  |  |  |  |
| --- | --- | --- | --- | --- | --- | --- |
|  | Second dose only | Aug 2021 | Sep 2021 | Oct 2021 | Nov 2021 | Dec 2021 |
|  | (N=54221) | (N=129868) | (N=142238) | (N=47515) | (N=8233) | (N=7190) |
| **Sex** |  |  |  |  |  |  |
| Female | 32363 (59.7%) | 71664 (55.2%) | 82343 (57.9%) | 29308 (61.7%) | 4959 (60.2%) | 4260 (59.2%) |
| Male | 21858 (40.3%) | 58204 (44.8%) | 59895 (42.1%) | 18207 (38.3%) | 3274 (39.8%) | 2930 (40.8%) |
| **Age Groups** |  |  |  |  |  |  |
| [16, 30) | 22633 (41.7%) | 1568 (1.2%) | 51574 (36.3%) | 18965 (39.9%) | 3428 (41.6%) | 3569 (49.6%) |
| [30, 40) | 12327 (22.7%) | 5889 (4.5%) | 37780 (26.6%) | 11873 (25.0%) | 2103 (25.5%) | 1593 (22.2%) |
| [40, 50) | 9458 (17.4%) | 25694 (19.8%) | 32911 (23.1%) | 8905 (18.7%) | 1365 (16.6%) | 1023 (14.2%) |
| [50, 60) | 5463 (10.1%) | 42065 (32.4%) | 13667 (9.6%) | 4835 (10.2%) | 662 (8.0%) | 547 (7.6%) |
| 60+ | 4340 (8.0%) | 54652 (42.1%) | 6306 (4.4%) | 2937 (6.2%) | 675 (8.2%) | 458 (6.4%) |
| **Socioeconomic status** |  |  |  |  |  |  |
| High | 12622 (23.3%) | 54111 (41.7%) | 58723 (41.3%) | 13315 (28.0%) | 2429 (29.5%) | 2312 (32.2%) |
| Middle | 29689 (54.8%) | 62546 (48.2%) | 69872 (49.1%) | 26750 (56.3%) | 4348 (52.8%) | 3712 (51.6%) |
| Low | 11831 (21.8%) | 12978 (10.0%) | 13468 (9.5%) | 7378 (15.5%) | 1444 (17.5%) | 1157 (16.1%) |
| Missing | 79 (0.1%) | 233 (0.2%) | 175 (0.1%) | 72 (0.2%) | 12 (0.1%) | 9 (0.1%) |
| **Social Sector** |  |  |  |  |  |  |
| General Jewish | 46446 (85.7%) | 123210 (94.9%) | 134380 (94.5%) | 43210 (90.9%) | 7352 (89.3%) | 6410 (89.2%) |
| Arab | 4243 (7.8%) | 2610 (2.0%) | 3904 (2.7%) | 2717 (5.7%) | 597 (7.3%) | 494 (6.9%) |
| Ultra-Orthodox Jew | 3532 (6.5%) | 4048 (3.1%) | 3954 (2.8%) | 1588 (3.3%) | 284 (3.4%) | 286 (4.0%) |
| **Cardiovascular diseases** |  |  |  |  |  |  |
| No | 51886 (95.7%) | 111880 (86.1%) | 137876 (96.9%) | 45952 (96.7%) | 7899 (95.9%) | 6936 (96.5%) |
| Yes | 2335 (4.3%) | 17988 (13.9%) | 4362 (3.1%) | 1563 (3.3%) | 334 (4.1%) | 254 (3.5%) |
| **Diabetes Mellitus** |  |  |  |  |  |  |
| No | 52315 (96.5%) | 110743 (85.3%) | 138150 (97.1%) | 46073 (97.0%) | 7940 (96.4%) | 6992 (97.2%) |
| Yes | 1906 (3.5%) | 19125 (14.7%) | 4088 (2.9%) | 1442 (3.0%) | 293 (3.6%) | 198 (2.8%) |
| **Hypertension** |  |  |  |  |  |  |
| No | 49678 (91.6%) | 87595 (67.4%) | 132847 (93.4%) | 43901 (92.4%) | 7538 (91.6%) | 6725 (93.5%) |
| Yes | 4543 (8.4%) | 42273 (32.6%) | 9391 (6.6%) | 3614 (7.6%) | 695 (8.4%) | 465 (6.5%) |
| **CKD** |  |  |  |  |  |  |
| No | 52108 (96.1%) | 106643 (82.1%) | 138241 (97.2%) | 45987 (96.8%) | 7888 (95.8%) | 6990 (97.2%) |
| Yes | 2113 (3.9%) | 23225 (17.9%) | 3997 (2.8%) | 1528 (3.2%) | 345 (4.2%) | 200 (2.8%) |
| **COPD** |  |  |  |  |  |  |
| No | 53860 (99.3%) | 125985 (97.0%) | 141682 (99.6%) | 47266 (99.5%) | 8176 (99.3%) | 7161 (99.6%) |
| Yes | 361 (0.7%) | 3883 (3.0%) | 556 (0.4%) | 249 (0.5%) | 57 (0.7%) | 29 (0.4%) |
| **Obesity (BMI ≥30)** |  |  |  |  |  |  |
| No | 47998 (88.5%) | 102774 (79.1%) | 125861 (88.5%) | 41762 (87.9%) | 7298 (88.6%) | 6465 (89.9%) |
| Yes | 6223 (11.5%) | 27094 (20.9%) | 16377 (11.5%) | 5753 (12.1%) | 935 (11.4%) | 725 (10.1%) |
| **Immunosuppression** |  |  |  |  |  |  |
| No | 52556 (96.9%) | 118570 (91.3%) | 138071 (97.1%) | 46233 (97.3%) | 7943 (96.5%) | 7000 (97.4%) |
| Yes | 1665 (3.1%) | 11298 (8.7%) | 4167 (2.9%) | 1282 (2.7%) | 290 (3.5%) | 190 (2.6%) |

CKD – Chronic Kidney Disease; COPD – Chronic Obstructive Pulmonary Disease.

The follow-up period exemplified Omicron’s rapid spread, with an increase in the daily numbers of infections and severe disease (**Figure 1**). During the examined Omicron-dominant period, a total of 101,737 booster breakthrough infections and 482 breakthrough infections resulting in COVID-19 hospitalizations or deaths were detected, of which 30,870 infections, 208 hospitalizations and 9 deaths were among those who received the booster dose early, in August 2021, whereas 1,082 infections, 4 hospitalizations and zero mortality were among the ‘newly vaccinated’ booster recipients, receiving their booster dose in December 2021. In that same Omicron period, 16,938 infections and 122 hospitalizations were recorded among those who received only two doses.

**Figure 1.**
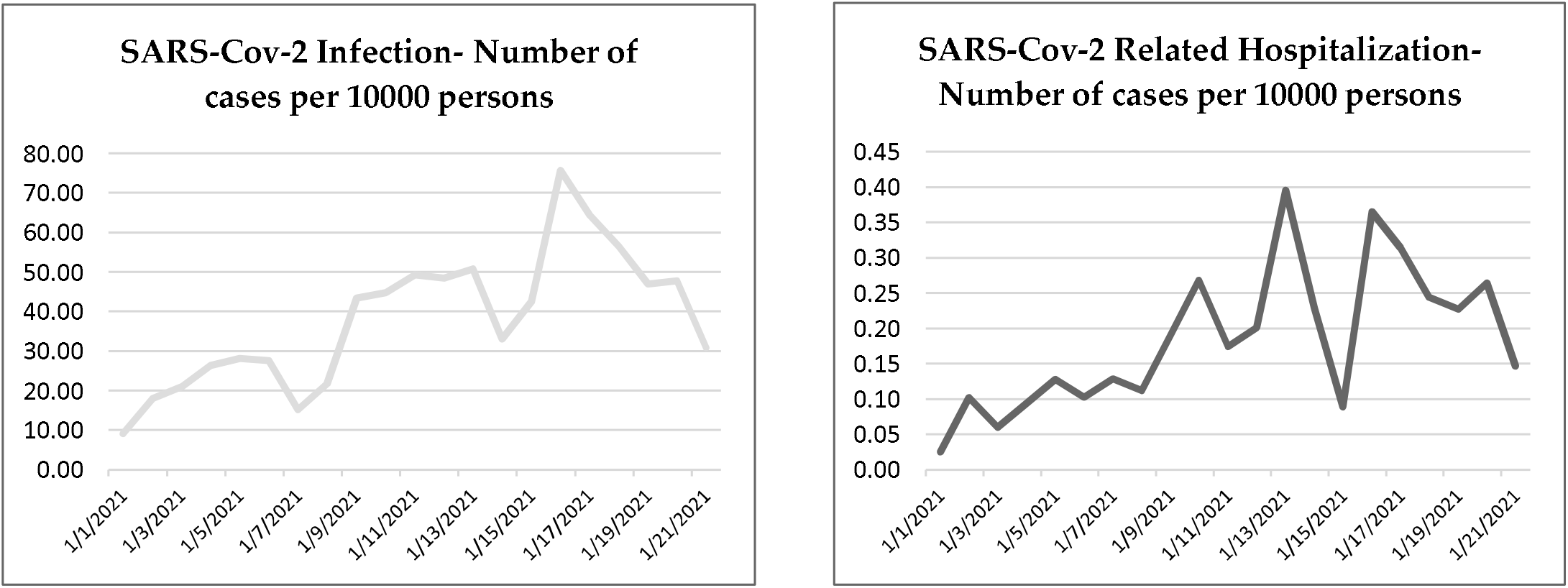
Daily SARS-Cov-2 infection and SARS-Cov-2-related hospitalization rates per 10,000 persons

### Main analysis – test-negative case-control design

**Table 2** details the results of the conditional regression in terms of the derived VE, whereas **table S1** details the adjusted Odds Ratio (aOR) in each month-from-vaccination. The aOR represents the ratio between the odds of testing positive or having severe COVID-19 during the January 1-21, 2022 period among those vaccinated in August 2021 (the first month of eligibility to the booster shot) and the respective odds in each subsequent month.

**Table 2.**
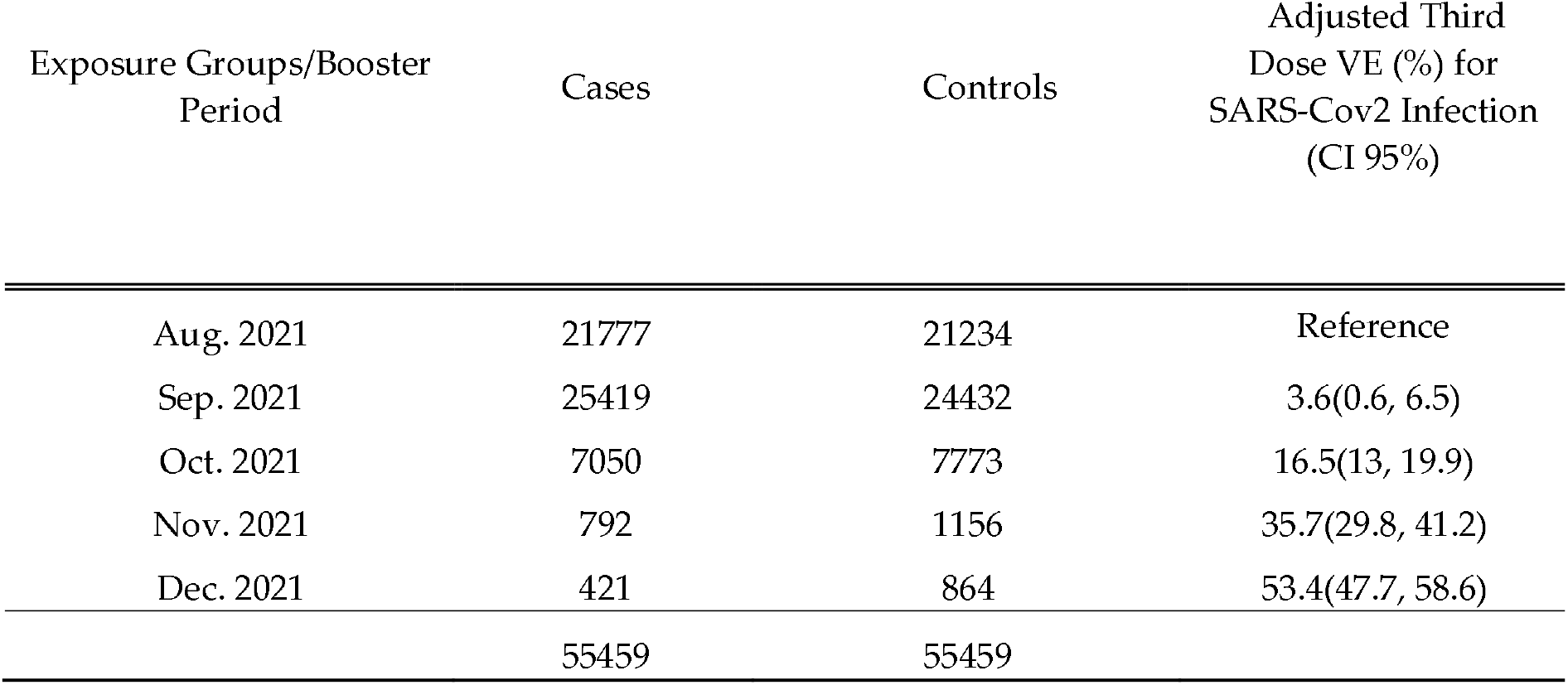
Main analysis; adjusted Third Dose Effectiveness (VE) against SARS-Cov2 infection. VE was defined as 100%*[1-(Odds Ratio)] for each of the month-since-vaccination categories. ORs were estimated with conditional logistic regression on a 1:1 matched dataset, and adjusted for comorbidities.

| Exposure Groups/Booster Period | Cases | Controls | Adjusted Third Dose VE (%) for SARS-Cov2 Infection (CI 95%) |
| --- | --- | --- | --- |
| Aug. 2021 | 21777 | 21234 | Reference |
| Sep. 2021 | 25419 | 24432 | 3.6(0.6, 6.5) |
| Oct. 2021 | 7050 | 7773 | 16.5(13, 19.9) |
| Nov. 2021 | 792 | 1156 | 35.7(29.8, 41.2) |
| Dec. 2021 | 421 | 864 | 53.4(47.7, 58.6) |
|  | 55459 | 55459 |  |

In the main analysis, we matched 110,918 cases and controls (in a 1:1 ratio) for the breakthrough infection analysis, and 562 cases and controls for the severe COVID-19 analysis (hospitalization or death). The vaccine effectiveness against infection of ‘newly’ vaccinated individuals (i.e. those receiving the booster in December, a month prior to the outcome period in January 2022) was 53.4% (95% Confidence Interval [CI], 47.7%-58.6%) compared to those vaccinated early, in August. VE declined rapidly in the subsequent two months, with 35.7% (95% CI, 29.8%-41.2%) VE for those vaccinated in November 2021 (two months prior to the outcome period) compared to August 2021, and 16.5% (%95 CI, 13.0%-19.9%) for the October 2021 vaccines. Those who received the booster shot in September 2021 had comparable odds of infection to those receiving the booster a month earlier, in August 2021 (**Table 2, Figure 2**). None of the comorbidities had a clinically significant effect **(Table S1**) on VE.

**Figure 2.**
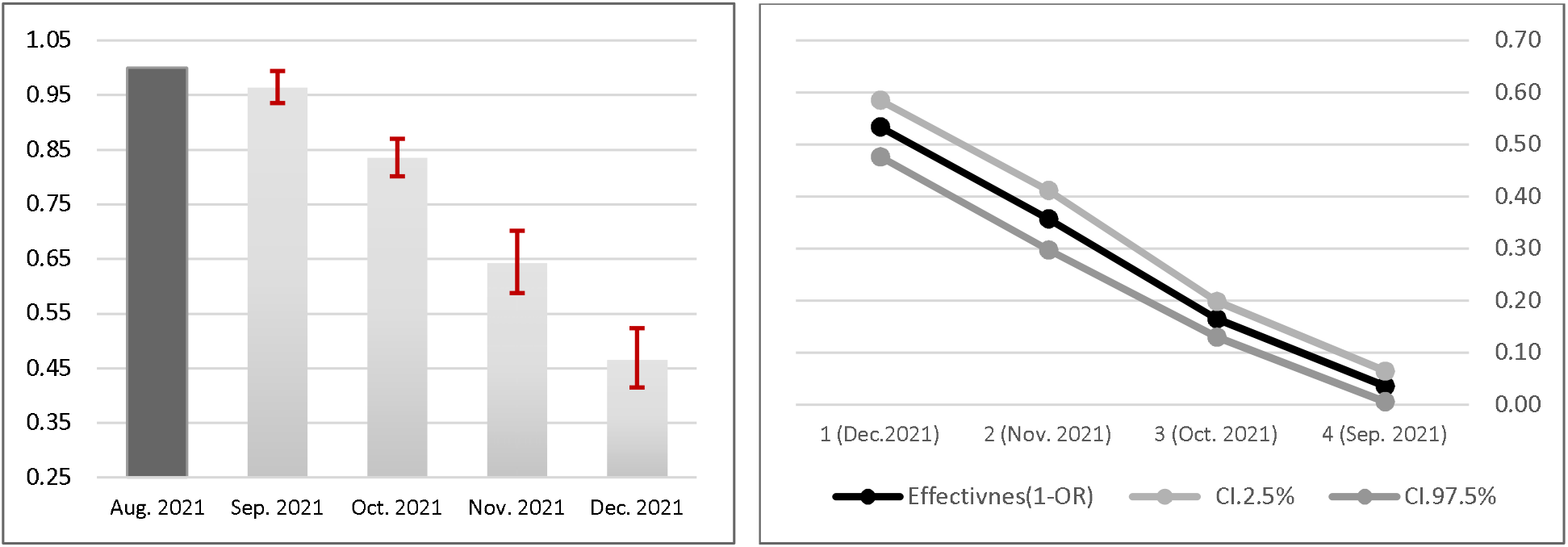
Adjusted Odds Ratio (left) and Vaccine Effectiveness (right) against SARS-CoV-2 breakthrough infection

As for severe disease, we did not reach significance implying a decreasing VE against hospitalizations or death as a function of time-since-inoculation (**Tables 2, S1**). However, the raw numbers are too small to support a reliable conclusion.

### Additional Analyses

The results of both sensitivity analyses (allowing for individuals to contribute multiple negative tests using a GEE logistic regression model and including tests performed on days 0-6 after the fourth vaccine dose) yielded estimates of VE that were overall similar to the results presented in the main analysis (**Tables S2 and S3** respectively).

The third additional analysis included changing the reference group; instead of a comparison within the booster recipients (that is, comparing the earlier vaccinees to late vaccinees), we compared each month of booster vaccinees to persons unvaccinated with the booster, namely those who received only two doses. Thus, we estimated the marginal effectiveness, or the added benefit of the booster compared to the second dose. In this analysis, we matched 128,854 persons for the breakthrough infection analysis and 722 persons for the severe COVID-19 analysis (hospitalization or death). The marginal effectiveness against infection of a booster dose given a month before the outcome period was at its peak at 59.4% (95% CI, 54.9%-63.5%). Effectiveness declined gradually with time from inoculation, reaching 16% (95% CI, 12.3%-19.5%) in those vaccinated 5 months prior to the outcome period compared to those not receiving the booster dose (Tables 3 and S4). As for the marginal effectiveness against severe disease, it seems that waning exists though to a much lesser degree, as effectiveness declines from 72.2% (95% CI, 37.8%-87.6%) 3 months after inoculation to 54.5% (95% CI, 13.4-76.1) five months after vaccination. However, numbers are small as also reflected by the confidence intervals.

**Table 3.** Additional analysis; adjusted Third Dose Effectiveness (VE) against SARS-Cov2 infection, compared to second-dose only vaccinees.

| Exposure Groups/Booster Period | Cases | Controls | Adjusted Third Dose VE (%) for SARS-Cov2 Infection (CI 95%) |
| --- | --- | --- | --- |
| Second Dose Only | 7866 | 6264 | Reference |
| Aug. 2021 | 21900 | 21588 | 16(12.3, 19.5) |
| Sep. 2021 | 25920 | 25623 | 18.3(15.2, 21.2) |
| Oct. 2021 | 7378 | 8583 | 29.1(26.1, 32) |
| Nov. 2021 | 872 | 1305 | 43.2(38.2, 47.8) |
| Dec. 2021 | 491 | 1064 | 59.4(54.9, 63.5) |
|  | 64427 | 64427 |  |

## Discussion

This research investigated the long-term duration of protection of the third (booster) dose of the BioNTech/Pfizer mRNA BNT162b2 vaccine against breakthrough infections largely due to the Omicron variant of SARS-CoV-2; a pivotal question in light of global discussions about the need for a fourth vaccine dose.

Our main analysis took a test-negative approach, comparing VE between recipients of the booster at different time points from inoculation. The analysis is based on the premise that had there been no waning of the booster protection over time, we would have seen no difference in infection odds at different times from vaccination (within the same calendrical outcome period). Nonetheless, we found that the effectiveness in each month since vaccination decreased significantly, whereas VE against infection compared to the first month of eligibility of the booster (August 2021) declined from 53.4% a month after vaccination to 16.5% three months after the booster dose. As for marginal protection of the third dose compared to the second dose (that is, protection gained on top of the one gained from two doses), we found a residual vaccine effectiveness of 16% after 5 months. Though the data suggests a milder waning of protection against severe disease, number of hospitalizations and mortality were too small to reach a reliable conclusion.

Studies have demonstrated that the third dose increases immunogenicity against SARS-CoV-2 as reflected by a rapid and broad immune response to the third BNT162b2 dose.^7,11^ Our results suggested a time dependent response with a primary increase in VE followed by waning of effectiveness. Overall, this waning of protection matches our knowledge of the course of the second dose of the BNT162b2, where protection increases immediately after a dose, but decreases rapidly within a few months. However, it seems that effectiveness could be declining faster than that of the second dose.^6,17^ The faster decline might partly be explain by an escape of the Omicron variant from neutralizing antibody responses,^18^ though further variant-specific evidence need to be established. Our results further point to the fact that waning of VE is not primarily affected by comorbidities, but rather that the most important factor in long-term VE is time from the last inoculation.

This study has some important limitations. The main limitation of every observational SARS-CoV-2 study is a potential bias stemming from healthcare related behavior as it pertains to PCR testing. This issue has been discussed in previous studies as well,^7,11,19^ and was more of a concern during the past months, in light of changing testing policies in Israel during the Omicron surge, where an overwhelming numbers of infected individuals did not allow the same test-for-all policy, implemented in previous waves. The test-negative design, which does not assume that a lack of a positive test equals a negative test and somewhat adjusts for testing volume, attempts to better mitigate this bias and is the reason we chose this method for this analysis., especially Additionally, the August 2021 vaccinees, who served as the comparison group in the main analysis were generally older and correspondingly sicker. This limitation has also been discussed in previous COVID research, as vaccination rollout generally prioritized chronically ill individuals. Nonetheless, our matching and adjustment approach renders residual confounding by indication less likely. Lastly, as the Omicron variant was the dominant variant in Israel during the analysis, the waning effect against other strains could not be evaluated. This could explain that in our secondary analysis of the marginal effectiveness, the additional benefit gained one month after inoculation was lower than the VE demonstrated in the same time frame in a previous research,^11^ investigating the Delta variant.

In conclusion, this study demonstrates significant waning of VE of the third dose of the BNT162b2 vaccine within a few months after administration. This waning effect, should prompt policy discussion as to vaccine development and future dose allocation.

## Supplementary Tables and Figures

**Table S1.** Conditional logistic regression results for SARS-Cov2 infection and COVID-19-related hospitalization.

|  | Infection |  | SARS-Cov2 Related Hospitalization |  |
| --- | --- | --- | --- | --- |
|  | OR | CI(95%) | OR | CI(95%) |
| Exposure Groups/Booster period(2) |  |  |  |  |
| Sep. 2021 | 0.964 | (0.935, 0.994) | 0.876 | (0.477, 1.607) |
| Oct. 2021 | 0.835 | (0.801, 0.87) | 0.812 | (0.345, 1.911) |
| Nov. 2021 | 0.643 | (0.588, 0.702) | 0.672 | (0.075, 6.052) |
| Dec. 2021 | 0.466 | (0.414, 0.523) | 2.681 | (0.321, 22.383) |
| Cardiovascular Diseases | 0.964 | (0.919, 1.012) | 1.611 | (0.928, 2.798) |
| Diabetes Mellitus | 0.945 | (0.903, 0.989) | 0.795 | (0.467, 1.354) |
| Hypertension | 0.994 | (0.96, 1.029) | 2.448 | (1.496, 4.005) |
| COPD | 0.890 | (0.808, 0.98) | 2.354 | (0.97, 5.71) |
| Immunosuppression | 0.949 | (0.901, 0.998) | 3.441 | (1.878, 6.305) |
| Obesity (BMI $\geq 30$) | 0.946 | (0.918, 0.976) | 0.915 | (0.566, 1.481) |

(2) Reference period: administration of booster during August 2021
COPD: Chronic Obstructive Pulmonary Disease

**Table S2.** GEE logistic regression results for SARS-Cov2 infection allowing for multiple negative tests

|  | Infection |  |
| --- | --- | --- |
|  | OR | CI(95%)(1) |
| Exposure Groups/Booster period |  |  |
| Sep. 2021(2) | 0.953 | (0.932, 0.974) |
| Oct. 2021(2) | 0.838 | (0.814, 0.863) |
| Nov. 2021(2) | 0.660 | (0.623, 0.7) |
| Dec. 2021(2) | 0.468 | (0.434, 0.505) |
| Sex(Male) | 1.097 | (1.08, 1.114) |
| Age Groups(3) |  |  |
| [30, 40) | 0.801 | (0.782, 0.821) |
| [40, 50) | 0.826 | (0.806, 0.846) |
| [50, 60) | 0.829 | (0.806, 0.852) |
| 60+ | 0.706 | (0.682, 0.73) |
| SES Group(4) |  |  |
| High | 1.006 | (0.99, 1.023) |
| Low | 0.932 | (0.906, 0.959) |
| Other | 0.779 | (0.628, 0.966) |
| Social Sector(5) |  |  |
| Arab | 1.047 | (0.998, 1.099) |
| Orthodox Jew | 2.435 | (2.334, 2.54) |
| Cardiovascular Diseases | 0.964 | (0.93, 0.999) |
| Diabetes Mellitus | 0.977 | (0.945, 1.011) |
| Hypertension | 0.958 | (0.934, 0.984) |
| COPD | 0.896 | (0.832, 0.966) |
| Immunosuppression | 0.929 | (0.894, 0.964) |
| Obesity (BMI $\geq 30$) | 0.936 | (0.916, 0.958) |
| Test Calendar Week(6) |  |  |
| Jan 8-14 | 2.393 | (2.345, 2.441) |
| Jan 15-21 | 3.496 | (3.427, 3.566) |
| Second Dose Month(7) |  |  |
| Feb. 2021 | 1.236 | (1.211, 1.261) |
| March 2021 | 1.270 | (1.238, 1.303) |
| Apr. 2021 | 1.041 | (0.98, 1.107) |
| May 2021 | 1.004 | (0.895, 1.127) |
| June 2021 | 1.035 | (0.854, 1.255) |
| July 2021 | 1.024 | (0.829, 1.265) |

(1) Robust 95% Wald confidence intervals were calculated using logistic generalized estimating equation (GEE) model, (3) Ref.: Age group [16, 30), (4) Ref.: High SES, (5) Ref.: Other, (6) Ref.: calendar week 01-07/01
COPD: Chronic Obstructive Pulmonary Disease

**Table S3.** Conditional logistic regression results for SARS-CoV-2 infection including PCR tests taken up to 6 days after the fourth vaccine dose

|  | Infection |  |
| --- | --- | --- |
|  | OR | CI(95%)(1) |
| Exposure Groups/Booster period(2) |  |  |
| Sep. 2021 | 0.974 | (0.945, 1.004) |
| Oct. 2021 | 0.846 | (0.813, 0.881) |
| Nov. 2021 | 0.690 | (0.631, 0.755) |
| Dec. 2021 | 0.471 | (0.419, 0.529) |
| Cardiovascular Diseases | 0.964 | (0.924, 1.006) |
| Diabetes Mellitus | 0.958 | (0.92, 0.998) |
| Hypertension | 0.988 | (0.957, 1.02) |
| COPD | 0.962 | (0.886, 1.045) |
| Immunosuppression | 0.908 | (0.867, 0.951) |
| Obesity (BMI $\geq 30$) | 0.952 | (0.925, 0.98) |

(1) 95% Wald confidence interval
(2) Reference period/exposure group: August 2021
COPD: Chronic Obstructive Pulmonary Disease

**Table S4.** Conditional logistic regression results for SARS-CoV-2 infection and COVID-19-related hospitalizations

|  | Infection |  | SARS-Cov2 Related Hospitalization |  |
| --- | --- | --- | --- | --- |
|  | OR | CI(95%)(1) | OR | CI(95%)(1) |
| Exposure Groups/Booster period(2) |  |  |  |  |
| Aug. 2021 | 0.840 | (0.805, 0.877) | 0.455 | (0.239, 0.866) |
| Sep. 2021 | 0.817 | (0.788, 0.848) | 0.450 | (0.237, 0.853) |
| Oct. 2021 | 0.709 | (0.68, 0.739) | 0.278 | (0.124, 0.622) |
| Nov. 2021 | 0.568 | (0.522, 0.618) | 0.508 | (0.095, 2.706) |
| Dec. 2021 | 0.406 | (0.365, 0.451) | 1.996 | (0.275, 14.468) |
| Cardiovascular diseases | 0.957 | (0.914, 1.003) | 2.454 | (1.496, 4.027) |
| Diabetes Mellitus | 0.950 | (0.909, 0.992) | 1.606 | (0.936, 2.756) |
| Hypertension | 1.000 | (0.967, 1.034) | 2.043 | (1.304, 3.2) |
| COPD | 0.910 | (0.828, 0.999) | 3.050 | (1.283, 7.251) |
| Immunosuppression | 0.937 | (0.892, 0.983) | 2.767 | (1.644, 4.656) |
| Obesity (BMI $\geq 30$) | 0.951 | (0.924, 0.979) | 0.763 | (0.484, 1.201) |

(1) 95% Wald confidence interval
(2) Reference period/exposure group: 2<sup>nd</sup> dose only
COPD: Chronic Obstructive Pulmonary Disease

